# Influence of time from symptom to treatment on quality of life in primary subclavian vein thrombosis

**DOI:** 10.1101/2025.03.19.25323979

**Authors:** Sara Blomstrand, Jonas Malmstedt

## Abstract

**Introduction:** Primary subclavian vein thrombosis (PSVT) is a rare (incidence 2/100,000) disease affecting predominantly young and physically active persons. A treatment protocol advocated by several studies includes catheter directed thrombolysis (CDT) within 14 days followed by decompressive surgery (TOD). A main purpose of this treatment is to prevent long term complications in form of Post-Thrombotic Syndrome (PTS), which manifests as reduced quality of life (QoL), assessed by the Disability of the Arm, Shoulder and Hand (DASH) score. CDT is rarely successful if started later than 14 days after the onset of symptoms. However, it is not known how a delay within the timespan of two weeks affects QoL in PSVT patients.

**Aims:** To investigate if there is a significant difference in QoL between PSVT patients receiving early treatment with CDT compared to those receiving delayed CDT.

**Material and Methods:** DASH score from 65 PSVT patients in Stockholm 2006-2018 was evaluated pretreatment and at one year follow up. Patients were divided into two groups depending on time from symptom to treatment with CDT. Early treatment group (0-3 days to CDT) was compared to delayed treatment group (4-14 days to CDT). Chi2 test was used to analyze differences in patient characteristics and Mann-Whitney-U test was used to identify any differences between the groups.

**Results:** No significant differences were detected (p=0.497)

**Conclusions:** No indication that treatment within 3 days from PSVT symptom onset is superior to treatment after 4-14 days was found. Because of the non-randomized design and small population, further studies are needed.

## Introduction

### Epidemiology

Upper extremity vein thrombosis (UEDVT) is a subgroup of deep vein thrombosis (DVT). Compared to lower extremity DVT patients, UEDVT patients are younger, leaner, and more likely to smoke (1). UEDVT in turn has two subgroups: primary and secondary UEDVT. Primary subclavian vein thrombosis (PSVT) is a rare disease, affecting predominantly young physically active persons without traditional risk factors for thrombosis. PSVT can be induced by strenuous activity of the arm such as competitive sports, music, or working with the arm in an elevated position (1,2). PSVT is less common than secondary UEDVT, which may be caused by malignancy, central venous catheters (CVC), pacemaker cables or thrombophilia (3). Predisposing factors for UEDVT besides CVC and malignancy are venous thromboembolism (VTE) heredity, hormone therapy and thrombophilia (such as Factor V Leiden mutation and prothrombin mutation) (3).

There is a lack of good quality modern data on incidence and predisposing factors for PSVT, as most studies either are not population based, or do not discriminate between primary and secondary thrombosis (3). The incidence of VTE (in all locations) is estimated to 104-183/100 000 (4). According to a Swedish study by Isma et al the incidence of UEDVT based on the Malmö population is 3,6/100 000(3). This study included 33 patients with PSVT corresponding to an estimated incidence of 2/100 000 (3) but this study excluded patients <18 years and had only 70% coverage.

### Pathogenesis

PSVT is caused by compression of the subclavian vein in the thoracic outlet due to anatomical variation. This is called thoracic outlet syndrome (TOS). In addition to venous thrombosis, TOS can give rise to arterial and/or neurological damage. In more than 50% of the cases PSVT is elicited by strenuous activities (called Paget Schroetter syndrome or effort-induced PSVT) (1,5). Typical activities inducing Paget Schroetter syndrome (PSS) include abduction and external rotation of the arm such as swimming, painting, or weight lifting (5). The pathogenic events of PSVT are micro trauma of the vessel wall caused by compression. The compression is caused by boney structures (clavicle, first rib or a cervical rib), ligaments (costoclavicular ligament), and muscles (anterior scalene muscle, subclavian muscle) (6). The micro trauma causes chronic inflammation, thickening and fibrosis of the vein. The intima is thereby damaged, which creates a thrombogenic surface (6).

### Clinical signs and diagnostics

Clinical signs of PSVT include pain, heaviness, swelling and discoloring of the arm, as well as distension of superficial veins of the arm and shoulder. The initial diagnostic tool is duplex ultrasound (sensitivity 78-100%, specificity 82-100%) (7,8). Contrast venography or computed tomography (CT) is used if suspicion of thrombosis remains after negative duplex ultrasound.

### Treatment

The aims of treatment are to halt the thrombus propagation, prevent recurrent episodes of DVT, prevent pulmonary embolism (PE), and to reduce the risk of post-thrombotic syndrome (PTS). If PSVT is diagnosed within 14 days from onset, catheter directed thrombolysis (CDT) combined with thoracic outlet decompressive surgery (TOD) of the vein is an optional treatment strategy (5,6). Active treatment with CDT and TOD is not recommended by the guidelines issued by the American College of Chest Physicians and from the American Heart Association, which states that thrombolysis and surgery shows no benefit compared to treatment with anticoagulation alone (9,10). On the contrary, CDT and surgery is recommended as a first line treatment by several centers (5,6,11,12). CDT is successful in 64-82% of the cases, and if started within 14 days from onset the success rate is reported to be nearly 100% (5,13). An unpublished pilot study of the Stockholm PSVT population suggests that patients treated with CDT within 14 days, followed by early TOD, had a lower DASH score at long term follow up than patients treated with anticoagulation alone, or with CDT and delayed TOD.

The frequency of PTS post PSVT is inconclusive, consequently the role of thrombolysis and surgery in treatment of PSVT is a field of debate (14,15). Additionally, there is a lack of randomized controlled studies evaluating the optimal treatment, mainly due to the low number of patients.

### Prognosis, complications and sequelae

Feared complications are recurrent thrombosis, pulmonary emboli (PE) and post-thrombotic syndrome (PTS). UEDVT patients seem to have a lower recurrence rate than lower limb DVT (3,16). But if the underlying anatomical problem is not corrected, one third of PSVT patients will have recurring clot and persisting symptoms (13). Pulmonary emboli is a potentially fatal complication to DVT. However, PE (incidence 117/100 000) (17) seems to be less common in UEDVT (2-35%) (18) than in lower limb DVT, and less common in primary (6%) than secondary UEDVT (13%) (19). Increasing PE incidence has a strong correlation with older age (4,17). Post-thrombotic syndrome (PTS) is a feared long-term complication of DVT. PTS stems from venous hypertension caused by outflow obstruction, in turn caused by incomplete recanalization or damaged vein valves (20). PTS manifests heterogeneously as signs and symptoms of chronic venous insufficiency (20). Symptoms of PTS include pain, swelling, heaviness and limited arm function. It occurs in 7-46% (15% mean) of patients with preceding UEDVT (21), and seems to be more common in PSVT patients (incidence as high as 75% has been reported) than in patients with secondary UEDVT (13,22). Incidence of PTS after UEDVT varies, in part due to lack of diagnostic standard for PTS (20).

### Measurement of Quality of life

Symptoms and disabilities in upper extremities, as well as outcome after treatment, can be measured as disease-specific Quality of Life (QoL) and is assessed by a questionnaire called the Disability of Arm, Shoulder and Hand (DASH) score (23,24). A Swedish adaptation of the DASH score, considered highly reliable (25), is used in this study. The DASH questionnaire consists of 30 questions with a five-response-option for each question. There are eight additional questions on work and leisure activities. The answers are converted into a scale from 0 to 100, 0 being equivalent to no symptoms, 100 to worst symptoms. According to a study on the American population, the DASH score in the general population is a means of 4 (26,27), with lower values correlating with a lower age. General population age group 19-34 years, which include mean and median age of PSVT patients, have a median DASH of 2 (IQR 0-6) (27). Minimal clinically important difference (MCID) in DASH is 13 points according to the DASH manual (27).

As one of the goals of PSVT treatment is to restore normal arm function, treatment outcome measured as QoL is essential, and the Swedish adaptation of DASH is a valid and reliable method of assessing QoL.

### Time aspects of treatment

As stated above, UEDVT and PSVT is often treated as DVT of lower extremities, though the etiologies differ (9,12,13,20,28). Furthermore, the diagnosis of PSVT is often delayed but the reasons for this delay (patient/doctor/misdiagnosis) are not well known. Patients with UEDVT usually present with symptoms within the first two or three days from onset of symptoms (29). Delayed diagnosis has potential impact on treatment options; CDT after more than two weeks from onset has poorer outcome than CDT started within two weeks (5,30). These studies use patency rate and symptom relief as outcome. Two studies report that CDT is more effective the younger the thrombus mass is, with a cutoff of 8 and 10 days respectively; the outcome was measured as symptom relief (31,32).

In summary, it is not known how a delay within the timespan of two weeks from symptom onset affects the outcome as QoL. It would be of clinical value to know how a delay within days affects long term outcome in QoL since it would contribute to the planning of patient treatment and to the decisions of in which order patient should be prioritized.

## Aim

The aim of this study was to compare Quality of Life (QoL) after a rapid and a delayed start of catheter directed thrombolysis (CDT) after first symptom, respectively.

## Materials and Methods

### Study design and study population

This was a retrospective cohort study including all PSVT patients treated with CDT in Stockholm county from March 2006 until March 2018. Patients were divided into two groups depending on time to treatment with CDT. The early treatment group was defined as having treatment initiated within 3 days from symptom onset. The delayed treatment group was defined as having treatment initiated 4-14 days after symptom onset.

QoL was measured as DASH score. The DASH score was assessed from questionnaires at two time points: at diagnosis i.e. before treatment, and at long term follow up. Long term follow up was defined as follow up after more than 100 days from diagnosis. Additionally, patients fill in the DASH score before and after any percutaneous transluminal angioplasty (PTA) or re-intervention. Patients generally filled in these questionnaires when visiting the hospital, otherwise the questionnaires were mailed to them. This is part of Sara Blomstrands Degree project 30 credits, spring 2018 at the Study Program in Medicine at Karolinska Institutet.

### Treatment protocol

The treatment strategy for this study population of PSVT patients with <14 days delay was to perform local CDT with Actilys. Actilys is a recombinant tissue plasminogen activator. Thrombolysis progression is controlled daily with a venogram. The purpose is to stop CDT when thrombus is resolved, or when resolution is no longer progressing. After successful CDT, patients are offered TOD to prevent recurrence of PSVT. Anticoagulation treatment (LMWH or Coumarin) is used for four weeks and antiplatelet therapy (usually acetylic-salicylic acid) is used for six months, given evaluations show that recovery is progressing as expected and there are no special circumstances (i.e. PE). Follow-up visits occur at 1, 3 and 6 months after hospital discharge and then yearly if all is well.

### Inclusion and exclusion criteria

PSVT was defined as duplex and/or CT-verified diagnosis of thrombosis in axillary and/or subclavian vein. Patients with secondary UEDVT (most commonly due to CVC, pacemaker or malignancy) and patients without sufficient medical records and/or without DASH data were excluded. Patients with recurrent or bilateral PSVT were only included once at their first PSVT. Inclusion and exclusion process is shown in Fig. 1.

**Figure 1.**
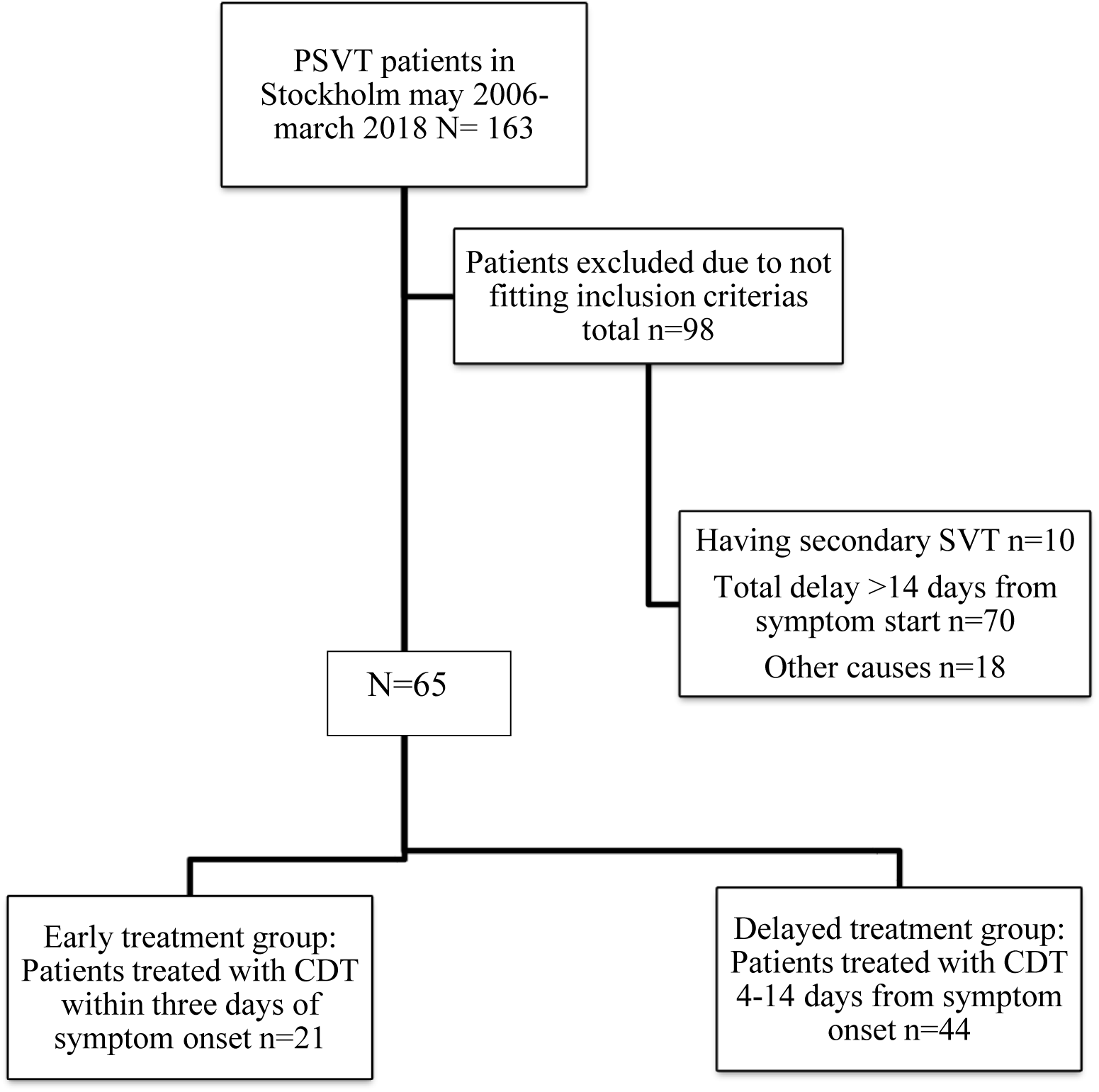
Inclusion and exclusion process. Causes of PSVT being secondary: Malignancy, pacemaker, clavicle trauma. Other causes for exclusion: Not having thrombosis (3), not having sufficient data in medical registries (5), not treated in Stockholm (4) other (6). PSVT=Primary Subclavian Vein Thrombosis, CDT=catheter directed thrombolysis.

### Outcome

The outcome was measured as disease specific quality of life (QoL) obtained from the DASH questionnaire. DASH was assessed before intervention and at late follow up (after >100 days). Comparison between the DASH score for patients in the two groups was presented as difference between groups and time points.

### Confounders and covariates

Potential confounders and covariates were patients’ sex, age, length, weight, smoking, use of anticoagulation or oral contraceptives before diagnosis, severity of initial symptoms, strenuous activity eliciting the thrombosis, work or sport affecting the arm, VTE heredity, previous VTE, thrombophilia, surgical intervention, events after surgery (e.g. PE, major bleeding, PTA), patients delay (time from symptom onset to health care contact) and doctors’ delay (time between health contact and treatment). Correlations between the suspected confounding factors and covariates, and the grouping and outcome were analyzed via a Chi^2^ test or a t-test.

### Type of data

PSVT patients’ data from medical records and DASH questionnaires was compiled into a database. Variables included: demographics; sex, age, smoking, length, weight, strenuous activity the last two weeks, use of oral contraceptives or anticoagulation, predisposing factors of PSVT including family history of VTE, previous VTE, thrombophilia; diagnostic imaging; time variables (time from debut of symptom to contact with health care, time from contact with health care to diagnosis, time from diagnosis to start of CDT or anticoagulation treatment); treatment details (length of CDT, surgery, type and duration of anticoagulation, complications, sequelae, follow-up with duplex ultrasound); DASH symptom score.

## Statistics

Continuous data were presented as means with ±standard deviation (SD) if normally distributed, and median with inter quartile range (IQR) if non-normally distributed. For demographics, differences in continuous and categorical variables were evaluated using a t-test and a Chi^2^-test respectively. DASH score was found to be skewed and therefore tested by non-parametrical tests. A two-way ANOVA was used in order to include sex and age as possible confounding factors to DASH score differences between the groups. We compared the DASH score pretreatment and at late follow up between the groups using a Mann-Whitney-U test. For evaluating change in the DASH score between pretreatment to late follow up in each group separately a Related-Samples Wilcoxon Sign Rank test was used. In order to compare the respective change in the DASH score between the groups a Mann-Whitney-U test was used. Data were collected via EpiData software, version 4.0.2.49 (Comprehensive Data Management and Basic Statistical Analysis System Odense. Denmark) and analyzed in IBM SPSS software version 24. A P-value < .05 was considered significant.

### Ethical considerations

Ethical permit was necessary since this study required reading of medical records and regional healthcare registries. This may be considered problematic from a personal integrity point of view, wherefore data in the study were anonymized. Results were statistically presented on group level. Patients were not exposed to any physical intervention nor physical risk. The benefit of getting indications to preferable treatment regime was considered greater than the inconvenience of any personal integrity compromise that the study may imply.

All patients treated for PSVT with surgery and/or CDT in Stockholm county are registered in SwedVasc (The Swedish National Quality Registry for Vascular Surgery) and are asked to fill in DASH-questionnaires. By receiving DASH-questionnaires they are informed about the study of which this report will be a part. They are informed that participating was voluntary, that data will be treated confidentially and without access from unauthorized staff, and that data will be presented anonymously. They are informed on how to access results of the study for which data is collected. Handing in the DASH-questionnaires was regarded as consent to participate in the study. Ethical permit was approved by the Regional Ethical Review Board in Stockholm 2017-01-18 (DNR 2016/2484-31/2).

## Results

### Population characteristics

Data from 163 patients treated for PSVT were collected. 65 patients were included in the study. Most of the excluded patients had either a delay of more than 14 days or causes that made their UEDVT secondary (Fig. 1). Patients were divided into two groups depending on time to treatment (early treatment =within 3 days, delayed treatment =4-14 days). Median age at time for diagnosis was 27 years (Table 1). 37 (52%) of the 65 patients were women, women were predominant in both groups. One patient in the early treatment group (5%) and six (14%) in the delayed group had no TOD, either because it was deemed unnecessary, or had low benefit expected. Baseline characteristics are presented in Table 1.

**Table 1.**
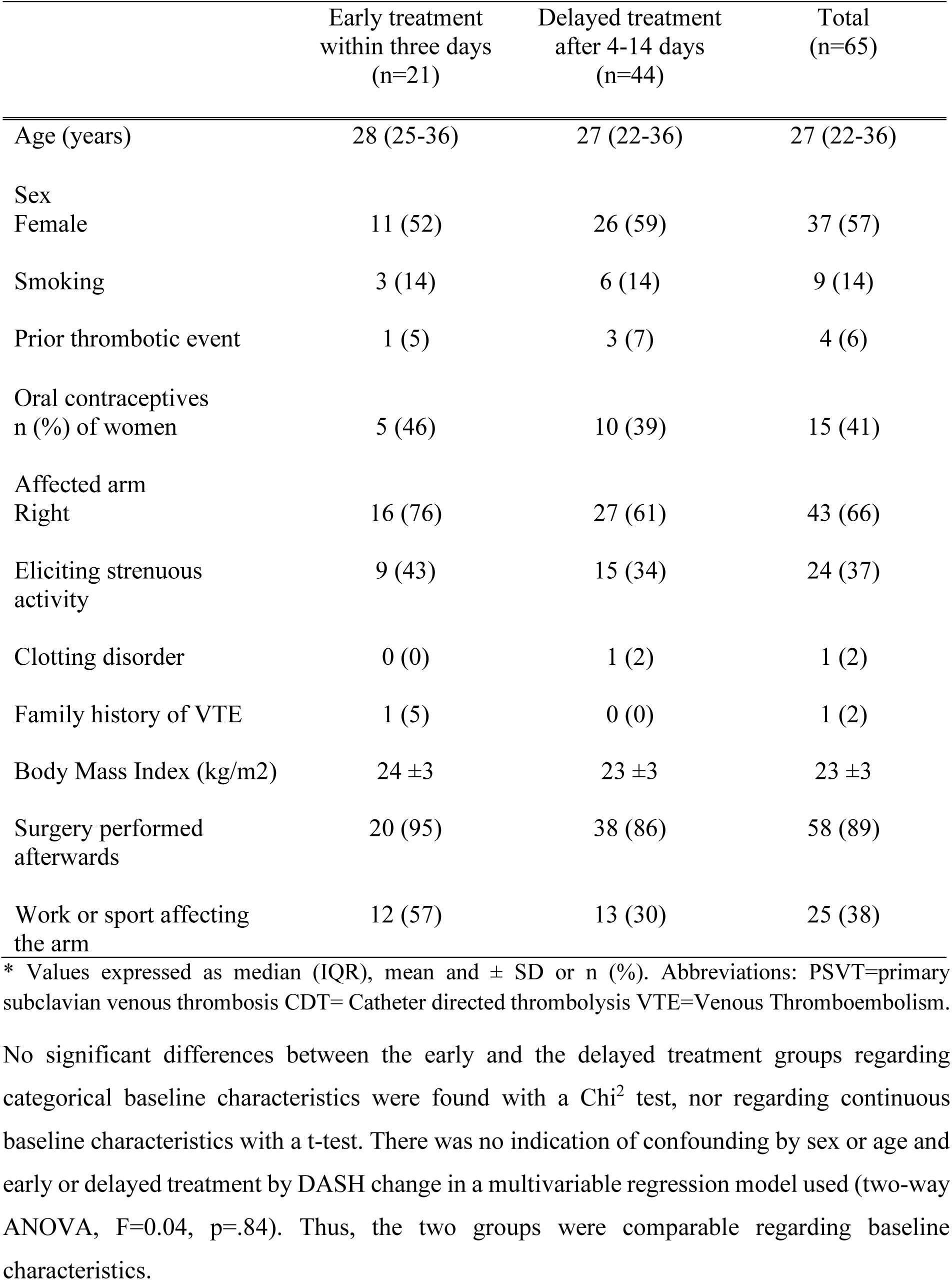
Patient characteristics and risk factors for thrombosis in 65 receiving early vs delayed treatment with CDT for PSVT. Patients are divided into two groups depending on their time from symptom to CDT treatment.

|  | Early treatment<br>within three days<br>(n=21) | Delayed treatment<br>after 4-14 days<br>(n=44) | Total<br>(n=65) |
| --- | --- | --- | --- |
| Age (years) | 28 (25-36) | 27 (22-36) | 27 (22-36) |
| Sex |  |  |  |
| Female | 11 (52) | 26 (59) | 37 (57) |
| Smoking | 3 (14) | 6 (14) | 9 (14) |
| Prior thrombotic event | 1 (5) | 3 (7) | 4 (6) |
| Oral contraceptives<br>n (%) of women | 5 (46) | 10 (39) | 15 (41) |
| Affected arm |  |  |  |
| Right | 16 (76) | 27 (61) | 43 (66) |
| Eliciting strenuous<br>activity | 9 (43) | 15 (34) | 24 (37) |
| Clotting disorder | 0 (0) | 1 (2) | 1 (2) |
| Family history of VTE | 1 (5) | 0 (0) | 1 (2) |
| Body Mass Index (kg/m<sup>2</sup>) | 24 ±3 | 23 ±3 | 23 ±3 |
| Surgery performed<br>afterwards | 20 (95) | 38 (86) | 58 (89) |
| Work or sport affecting<br>the arm | 12 (57) | 13 (30) | 25 (38) |

\* Values expressed as median (IQR), mean and ± SD or n (%). Abbreviations: PSVT=primary subclavian venous thrombosis CDT= Catheter directed thrombolysis VTE=Venous Thromboembolism.

No significant differences between the early and the delayed treatment groups regarding categorical baseline characteristics were found with a Chi^2^ test, nor regarding continuous baseline characteristics with a t-test. There was no indication of confounding by sex or age and early or delayed treatment by DASH change in a multivariable regression model used (two-way ANOVA, F=0.04, p=.84). Thus, the two groups were comparable regarding baseline characteristics.

The delay in time to treatment could be explained partly by patients delay and partly by delayed initiation of treatment. Doctors delay did not contribute to the total time to treatment. (Table 2).

**Table 2.** Time aspects of delay in 65 patients treated for PSVT. Patients divided into two groups depending on time from symptom start to CDT treatment. Values expressed as median (IQR).

|  | Early treatment<br>within three days<br>(n=21) | Delayed treatment<br>after 4-14 days<br>(n=44) | Total<br>(n=65) |
| --- | --- | --- | --- |
| Patient delay (days) | 0.0 (0.0-1.0) | 3.0 (1.0-6.5) | 1.0 (0.0-4.0) |
| Doctors delay<br>(days) | 0.0 (0.0-0.5) | 0.0 (0.0-3.0) | 0.0 (0.0-1.0) |
| Time to diagnosis<br>(days) | 1.0 (0.0-1.0) | 4.5 (3.0-7.0) | 3.0 (1.0-5.5) |
| Time to start of<br>CDT (days) | 2.0 (1.0-2.0) | 6.5 (5.0-9.8) | 5.0 (2.0-9.0) |

Abbreviations: PSVT=primary subclavian venous thrombosis CDT= Catheter directed thrombolysis

### Associations between the DASH score and time

#### The DASH score at baseline and long-term follow up

We found no significant difference in the patient-reported primary outcome (median DASH score) at late follow up between the early and delayed treatment groups. Both groups had comparable baseline DASH scores (Table 3).

**Table 3.** DASH median at baseline and long term follow up. Patients were treated with CDT and anticoagulation within three days from symptom onset or after 4-14 days from symptom onset.

|  | Early treatment<br>(n= 21) | Delayed<br>treatment (n=44) | Total<br>(n=65) | P** |
| --- | --- | --- | --- | --- |
| DASH at<br>baseline *(n=43) | 28.9 (11.7-43.1) | 21.7 (13.3-48.3) | 28.7 (13.3-47.5) | 0.880 |
| DASH at long<br>term follow up<br>*(n=40) | 4.2 (0-9.4) | 1.7 (0-5.2) | 2.5 (0.0-7.1) | 0.497 |

\*Values expressed as median (IQR). \*\* Mann-Whitney-U test. Abbreviations: DASH = Disability of the Arm, Shoulder and Hand. AK=anticoagulants. CDT= catheter directed thrombolysis.

#### Change in the DASH score from pretreatment to long-term follow up

Out of the 65 patients, 25 (early treatment group n=9, delayed treatment group n=16) had submitted a DASH score both at pre-intervention and at long term follow up and were thus eligible for analysis of change in DASH. No significant difference between the groups respective change in DASH could be detected by a Mann-Whitney-U test (p 0.692) (Table 4).

**Table 4.** Change in the DASH from baseline to long term follow up. Change measured in 25 patients treated with CDT within three days from symptom onset (early treatment) or after 4-14 days from symptom onset (delayed treatment).

|  | Early treatment<br>*(n=9) | Delayed<br>treatment<br>*(n=16) | Total<br>*(n=25) | P** |
| --- | --- | --- | --- | --- |
| Change in<br>DASH score | -15.3 ± 12.4 | -21.5 ± 21.0 | -19.3 ± 18.4 | 0.692 |

\*Values expressed as negative mean ( ± SD) \*\* Mann-Whitney-U test. Abbreviations: DASH = Disability of the Arm Shoulder and Hand. CDT= Catheter Directed Thrombolysis

Both the early and the delayed group’s DASH scores showed significant improvement from pretreatment to long-term follow up, as assessed with a non-parametric test (p = .012 resp. p = .001, Related-Samples Wilcoxon Sign Rank test). Both treatment groups showed improved QoL at late follow up, with median 2.5 (IQR 0.0-7.1) and mean 2.8 (±4.4) DASH score which is close to the healthy population mean DASH score of 2.0 in age group 19-34 years (Fig. 2).

**Figure 2.**
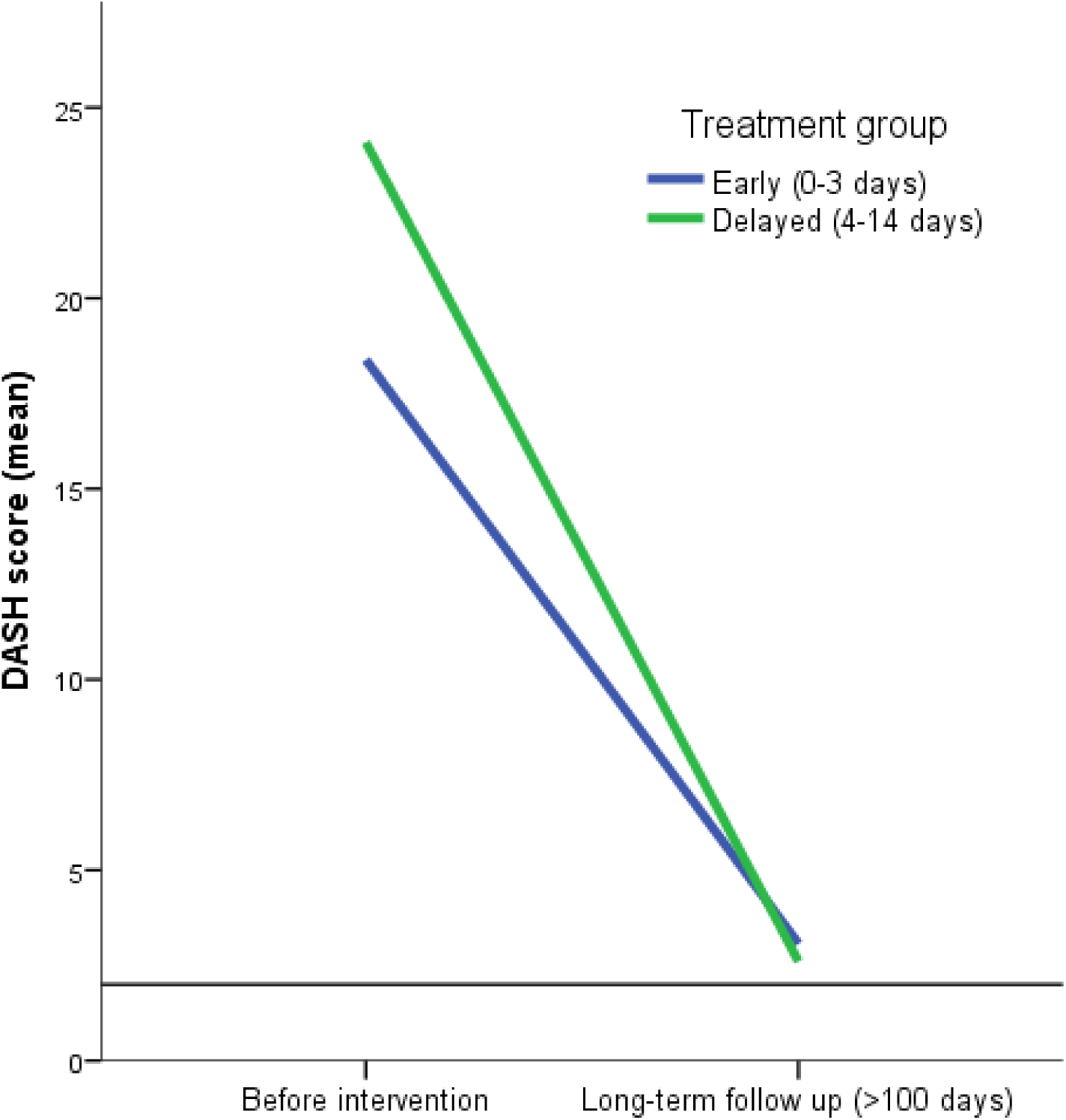
Mean DASH score pretreatment and at long term follow up. DASH was measured in PSVT patients receiving early (within 3 days from symptom start) vs delayed (4-14 days after symptom start) treatment with CDT. Mean DASH (2) in normal healthy population age 19-34 indicated in black. Abbreviations: PSVT= Primary subclavian vein thrombosis. DASH = Disability of the Arm Shoulder and Hand. CDT= Catheter Directed Thrombolysis

## Discussion

This retrospective cohort study of 65 patients with PSVT treated in Stockholm May 2006-March 2018 aims to compare quality of life (QoL) differences between patients with early or delayed treatment with catheter directed thrombolysis (CDT). QoL was measured in a questionnaire as the extent of disability of the arm shoulder and hand (DASH) score. Differences in time from symptom onset and treatment is compared between patients treated within 3 days (early treatment) and after 4-14 days (delayed treatment) respectively. The DASH score was assessed at pretreatment and at long term (>100 days) follow up. Patients treated early showed no significant difference in long term DASH score compared to patients with delayed treatment. Nor did the change in DASH score from pretreatment to late follow up differ significantly between the groups.

Median age was 27 (range 22-36), which is in the lower range compared to previous results (5,33). 57% were women, consistent with some studies (1,3) but a higher percent than in other studies (5,28). This difference may be due to a higher percent of female athletes in recent years (34). Some 37% reported strenuous activity prior to the onset. The numbers previously reported range from 25-80% of PSVT (5,30,31,35). Data of frequency of strenuous activity prior to PSVT might be less definite than data of for example age and sex, due to patient recall bias and the need for interpretation of medical records to assess the frequency.

A few studies have investigated time to treatment as a predictor of outcome after CDT with following TOD. Consensus seems to be that treatment started within two weeks from the onset of symptoms have a better outcome than CDT started later than two weeks from the onset of symptoms (5,30–32). There is data presented as secondary outcomes in some studies: Two studies by Zimmerman et al (32) and Adelmann et al (31) report CDT to be more effective the younger the thrombus mass is: Zimmerman et al reported 82% success of CDT in 11 patients treated within 8 days, defined as patency of the vein (32). In seven patients treated after >10 days (12-180 days), none were successful. Follow-up time was six months, and frequency of TOD was not reported. Adelmann et al reported success in 79% of 17 patients treated within 8 days. Three patients were treated later, only one of them had success (31). Success was defined as vein patency, but Adelmann et al also reports lack of symptoms in all those with successful CDT. The definition of symptoms was discoloration, edema, discomfort or lack of mobility in rest or abduction of the arm. Wilson et al (36) reported 100% recanalization in patients treated with CDT within one week (total recanalization in five and partial recanalization in two). Patients had no symptom of PTS or swelling at follow up, up to five years after treatment. No patients treated later than one week after onset were part of the study, and patients did not have TOD. Doyle et al (30) reported success measured as restored flow in 100% of 16 PSVT patients treated with CDT and anticoagulation within 14 days from symptom onset. In the same study four patients were treated with CDT later than 14 days, all four CDT attempts failed (30). All patients (except two in the delayed group) later had TOD. At the five years follow up the same study reported symptom relief and patency rate of approximately 85% similarly in both groups. The similarity of success at long term between the two groups might be related to a lower symptom score in the delayed group at start, as baseline symptom score was not reported. Additionally, a reason for high symptom relief at long term in PSVT patients treated with CDT (later than 14 days), has been suggested to be that collateral circulation might begin to relieve the subclavian vein circulation, thereby not making the clot resolution by CDT as crucial for symptom improvement, or at least contributing to the effect of any clot resolution in the subclavian vein (37). These four studies were studies on a limited material. In a larger study Molina et al reported 100% recanalization after CDT and anticoagulation within 14 days in 97 patients (33). All patients were offered TOD after CDT and anticoagulants treatment, and at long term follow up after mean 5.2 year, 100% still had patent veins. None of the five above-mentioned studies had any delayed group treated within 14 days to contrast to the early treatment. Furthermore, patient-related outcomes were only reported as subjective dichotomous variables (e.g. improved/not improved, asymptomatic/symptomatic). It is therefore difficult to compare these results with ours, even though measurement of subjective absence of symptom is a part of what the DASH score sets out to quantify, in a reproducible manner.

In a larger retrospective study by Urschel et al, 506 patients received CDT and 96% of them improved significantly (13). In this study, similar to our own, outcome as symptom relief was included. Here improvement was based on three categories: pain relief, employment and limited recreation. A difference depending on time to treatment was reported, but the cutoff was set to six weeks. Out of 42 patients treated later than six weeks after onset of symptoms, only 24 were categorized as having excellent to good results (13). The low success rate of CDT after six weeks delay is not unexpected, since patients treated in a large time span after 14 days are included. It is rather the good outcome of 96% success in patients in such a large time span as six weeks that could be perceived as surprising. But since the distribution of patients’ time to treatment is not presented further than earlier or later than six weeks from symptom start, it is not certain that time to treatment <14 days correlates with better outcome in the Urshel study.

To summarize, our results are not directly comparable to earlier studies on time impact on success of CDT. Firstly, outcome is often dichotomized as successful or not. Secondly, previous studies have focused on patency of the vein whereas we measured QoL. Where symptom burden has been assessed, a standardized model such as the DASH score has not been used, and symptom at baseline has not always been reported. Thirdly, early or delayed treatment has been measured as within eight days to six weeks, none have contrasted their early group to a delayed group that is treated within a time span as early as ours. In addition, our follow-up time was longer compared to some of the earlier studies. It would be unfortunate to assess outcome in terms of QoL, or even symptom relief, too early after PSVT, since the treatment following CDT often include surgery and physical activity, which may take several months to show results.

If we still are to compare our results to the ones mentioned above, we can question why no difference was seen between patients receiving early vs delayed treatment in our study, when it is reported that early treatment is superior in the studies earlier discussed. One hypothesis is that there is a true difference in patency and symptom relief as defined in the mentioned studies, but no difference in QoL. Another hypothesis is that there was a difference in symptom relief at earlier time points, but at late follow up which we define as >100 days (and often was at one year follow up), differences are no longer present. Another possibility is that neither the studies above had any difference between patients treated within three days and patients treated 4-14 days. This is not possible to tell from existing material. The treatment following CDT must also be considered to do a fair comparison.

We found no evidence that early treatment within three days is favorable compared to delayed treatment after 4-14 days, in terms of quality of life measured as DASH score. Our results need to be confirmed in a larger population with prospective data before recommendations to change current clinical practice to accepting delayed treatment.

The median DASH score from pre-treatment and the median DASH score at late follow up differ significantly for each group respectively. The minimal clinically important MCID of 13 points between pre-intervention and follow up (27,38) was detected in both groups: Mean 15.3 points in the early treatment group and 21.5 points in the delayed treatment group. These results are consistent with previous studies arguing that treatment with CDT within 14 days has a superior outcome compared to conservative treatment with anticoagulant medication in PSVT patients. Furthermore, the median DASH score at late follow up of 2.5 is lower than mean DASH healthy population, which is reported to have a mean DASH score of 4, and is close to the healthy population mean of 2 for age group 19-34 years (26,27). These results might be related to the healthy physically active lifestyle that PSVT patients often lead prior to their onset.

### Strengths and limitations

To our knowledge, this is the first study evaluating outcome after different time spans between symptom onset and CDT treatment, using disease specific QoL. It is also the first attempt to examine effects of delayed CDT on outcome, within the timeframe of 14 days. The generalizability benefits from our cohort being population based and not including tertial referrals.

A limitation of this study was that not every patient has answered a DASH-questionnaire both before intervention and at long time follow up. This made the already limited number of patients eligible for analysis even smaller. The reason was in part that some patients in the study are diagnosed less than a year ago and have not yet had a late follow up visit. The small number of patients, due to the low incidence rate, and that not all patients get to health care within 14 days, was a limiting factor which was handled by including patients from over a decade.

Our study does not include clot burden. It could be argued that the clot burden and time correlate. A greater clot burden has been shown to correlate negatively with success of CDT (11). If time to treatment correlates to long term DASH score, the true predictor could be clot burden instead of time. However, this would not necessarily call for altered treatment strategies.

The manual collecting of data from medical registries is another limiting factor because manual errors may occur. This was partly adjusted for by using EpiData, which was programmed to only allow certain values, limiting the risk of erroneous data entries, and to make certain fields of data entry mandatory. This minimized the risk of overlooking crucial key points in data collection from each medical record.

### Clinical applications

It is essential to know how to prioritize patients to surgery. The study design choice of a delay of over four days has a clinical implication in forms of giving information on whether a CDT could be postponed over the weekend without undesired effects on the outcome or not. Our results indicate that start of CDT may be delayed over a weekend without a negative impact on outcome. Median time to CDT treatment is 5.0 days, consequently, by our definition, most patients receive delayed treatment. Time to diagnosis is shorter, median 3.0 days. The discrepancy between time to diagnosis and time to treatment with CDT might imply that there is a possibility to treat patients even earlier with CDT. According to our results, there is no indication of need of urgent start of CDT.

### Future studies

Further studies, including a randomized design for comparison of early or delayed treatment, would be of value. It would be interesting to additionally include a measure of clot burden and degree of adjusted collateral vessel circulation, to analyze if this has impact on outcome after CDT and anticoagulants treatment. It could be argued that the clot burden and adjustment in form of circulation through collateral vessels is a reason for different outcomes of anticoagulants and CDT, correlating with time from symptom to treatment. To study this, a multivariate model including assessments of clot burden and collaterals vessels at first visit could be used. Data from venograms performed at first visit would be of use to provide measurements of size and degree of flow obstruction, as well as the detailed data of symptom characteristics which are assessed from the DASH questionnaire.

It would also be interesting to study reasons for delay, even though delayed treatment was not found to be an indication of inferior outcome in this study. Many PSVT patients seek health care later than 14 days from symptom start and are therefore not eligible for CDT treatment. The remaining option for them is conservative treatment, and delayed surgery is indicated in case of severe persisting symptoms. Delayed surgery has been associated with inferior outcome in QoL. It would be of clinical value to find the main reason for delayed prognosis to shorten the time to treatment.

## Conclusions

The aim of this study was to investigate if there was a significant difference in disease specific QoL between patients receiving early vs delayed CDT for PSVT. It cannot be concluded that a more urgent treatment is favorable, compared to a treatment within 4-14 days. To confirm these results there is a need for further prospective studies with randomization between early vs delayed treatment, and on a more numerous population.

## Data Availability

Individual participant data underlying the results reported in this Article will be made available after de-identification, alongside a data dictionary, study protocol, and informed consent form. The data will be available at Article publication and for 10 years subsequently. Data will be shared for individual participant data meta-analysis with other members of the research community who have an affiliation to a recognised medical university. Data will only be shared with investigator support and after approval of a proposal, and with a signed data access agreement. Additional restrictions apply according to Swedish law.

## Acknowledgements

I would like to thank my supervisor Jonas Malmstedt for his patient guidance and support. Data collection was done in collaboration with future colleague Magdalena Lublin, her companionship through this project has been of great value. Additionally, I want to thank my coordinator Vladimer Darsalia and my opponents for their valuable comments.

## Abbreviations

CDT: Catheter directed Thrombolysis
CT: Computed Tomography
CVC: Central Venous Catheter
DASH: Disability of Arm Shoulder and Hand
DVT: Deep Vein Thrombosis
LEDVT: Lower Extremity Deep Vein Thrombosis
LMWH: Low Molecular Weight Heparin
MCID: Minimal Clinically Important Difference
PE: Pulmonary Emboli
PSS: Paget Schroetter Syndrome
PSVT: Primary Subclavian Vein Thrombosis
PTA: Percutaneous Transluminal Angioplasty
PTS: Post-Thrombotic Syndrome
SVT: Subclavian Vein Thrombosis
TOD: Thoracic Outlet Decompression
TOS: Thoracic Outlet Syndrome
UEDVT: Upper Extremity Deep Vein Thrombosis
VTE: Venous Thromboembolism
QoL: Quality of Life

